# Characterizing the cascades of care experienced by pregnant adolescents in three African countries; Ethiopia, Kenya and South Africa: A Cohort Study

**DOI:** 10.1101/2025.05.16.25327748

**Authors:** Beatrice Amboko, Irene Mugenya, Emily Odipo, Rose Jepchumba Kosgei, Wen-Chien Yang, Shalom Sabwa, Gebeyaw Molla Wondim, Nompumelelo Gloria Mfeka-Nkabinde, Londiwe Mthethwa, Nokuzola Cynthia Mzolo, Margaret E. Kruk, Catherine Arsenault, Jacinta Nzinga

## Abstract

The capacity of health systems to provide quality care for pregnant adolescents remains underexplored. This study examined the quality of care and adverse delivery outcomes (mistreatment and obstetric complications) among pregnant adolescents in Ethiopia, Kenya, and South Africa using data from the MNH eCohort longitudinal survey. This study followed 3,051 pregnant women from their first antenatal care (ANC) visit to postpartum period. We used descriptive analysis to compare outcomes between adolescents (<20) and adults (20+), and logistic regression to identify factors associated with mistreatment and adverse delivery outcomes among adolescents. A total of 380 adolescents (mean age 18) and 2,671 adults (mean age 28) were included in the analysis. Adolescents were more likely to be unmarried (63%), particularly in Kenya (53%) and South Africa (99%), while most in Ethiopia were married (95%). Education level varied, with many Ethiopian adolescents lacking formal education. Only 23% of adolescents attended their first ANC visit in the first trimester compared to 30% of adults (p=0.003), with lower follow-up testing and adherence to iron and folic acid supplementation. The mean ANC visits was 3.8, with higher levels among adults in Ethiopia and Kenya. Over 95% of women delivered in health facilities, but consent for vaginal examinations was low (46%). Mistreatment was reported by 20% of women, with higher rates among adolescents (27% vs. 19%, p=0.003). A higher risk of obstetric complications was associated with rural residence (OR: 2.25, 95% CI: 1.09–4.63) and antenatal depression (OR: 2.43, 95% CI: 1.19–5.00). Mistreatment was associated with rural residence, public facility visits, and experiencing intimate partner violence. Kenyan adolescents, privacy during delivery, and high quality of care rating care were protective factors for mistreatment. Adolescents face critical gaps in maternal healthcare, requiring strengthened adolescent-friendly services, rural healthcare investments, and respectful maternity care policies to improve outcomes.

## INTRODUCTION

Adolescent pregnancy is a global issue, causing physical, psychological and socioeconomic challenges for young mothers, newborns and their communities (1, 2). Adolescence is the period between 10 and 19 years of age (3). This is an especially vulnerable period as it represents the period when one is transitioning to adulthood and is characterised by physical, psychological, and emotional developmental changes (3, 4). Each year, an estimated 21 million pregnancies occur among adolescents in low and middle-income countries, with a majority of the cases taking place in sub-Saharan Africa (SSA) (2, 5, 6). Half of the pregnancies among adolescents are unintended, putting them at increased risk of pregnancy-related complications, including pre-eclampsia, anaemia, postpartum haemorrhage, obstructed labour, postpartum depression and sepsis and acquiring HIV or other sexually transmitted infections (5, 7). Within the SSA region, adolescents from socio-economically disadvantaged backgrounds experience higher pregnancy rates and poorer outcomes disproportionately affect adolescents from socio-economically disadvantaged backgrounds and with lower levels of education, hence the need to address this public health issue (8, 9).

Furthermore, infants born to adolescent mothers face an elevated risk of complications or death, including stillbirth, low birth weight, and premature births (10, 11). In addition to the physical risks, adolescent pregnancies also have adverse psychosocial and economic consequences, such as school dropout, social stigma (which is less common among their male counterparts), early marriages and increased vulnerability to abuse (12, 13). These adverse effects can largely be attributed to the age-related and developmental processes occurring during this period, their dependency on others and unequal power dynamics, as well as the delayed seeking of maternal health care services until the pregnancy is well advanced (14, 15).

The health system should begin by helping adolescents delay the age of their first pregnancy. However, when adolescents become pregnant and choose to carry the pregnancy to term, health systems must ensure that they have access to high-quality and respectful health services that are tailored to their needs and preferences (9). Previous studies have shown that adolescents are less likely to receive a minimum number of antenatal care (ANC) consultations and to give birth in health facilities (16, 17).

Additionally, among those who access these services as recommended, it has been noted that they are at risk of receiving lower quality care and social support than adult women seeking the same services (18). This shows that persistent gaps hinder access to quality maternal and neonatal health services among adolescents. Nonetheless, evidence on the quality of maternal health services for adolescents in SSA remains limited. This study examined maternal healthcare quality and continuity, as well as the factors associated with mistreatment and obstetric complications among adolescents in Ethiopia, Kenya, and South Africa.

## STUDY METHODS

### Data source

This analysis uses data from the Maternal and Newborn Health (MNH) eCohort in Ethiopia, Kenya and South Africa, a longitudinal health system quality study that enrolled women face-to-face during their first ANC visit and followed them over the phone through pregnancy, delivery, and the postnatal care period. Detailed information on the data collection methods is published elsewhere (19, 20). In summary, pregnant women aged 15 years and above were enrolled in the study after their first ANC visit. The women were enrolled from both public and private health facilities in Ethiopia and Kenya and public facilities only in South Africa. Data collection methods included a baseline face-to-face interview at the health facility and repeated phone interviews during the antenatal period and after delivery. Information on demographic characteristics, content of care (assessments, tests, treatments, counselling), and perceptions of the quality of care received was collected. Data were collected between April 2023 and February 2024 in Ethiopia, June 2023 and July 2024 in Kenya, and April 2023 and August 2024 in South Africa. In this manuscript, we present data collected from adolescent girls aged 15-19 years and adult women aged 20+ years recruited into the study at the first ANC visit until 6 weeks post-delivery.

### Measures

#### Quality and continuity of care

To understand the quality of care provided, we assessed both the content of care and the participant’s perceptions of the services received during the first ANC visit, follow-up ANC visits, delivery care and the early postnatal period. Specifically, we examined the assessments, tests and treatments they received, including the recommended supplements during pregnancy. Box 1 outlines the specific content of care indicators for each service across the maternal healthcare continuum.

##### Box 1.

Measures of quality and continuity of care across the maternal healthcare continuum

###### First ANC visit

1. First ANC attendance in the first trimester
2. First ANC screening and treatment completeness index^a^
3. First ANC counselling completeness index^b^
4. Overall rating of the quality of first ANC visits as very good/excellent^c^

###### Follow-up (FU) ANC visits

1. The proportion with at least one blood test after the first ANC visit
2. The proportion with at least one urine test after the first ANC visit
3. The proportion who received food supplementation at least once during antenatal period among all women
4. The proportion who had ultrasound done at one point (baseline or FU ANC visits)
5. Adherence to iron and folic acid supplementation (IFAS)^d^
6. The proportion who were counselled on postpartum family planning at least once
7. The number of additional and total ANC visits

###### Delivery care

1. The proportion who delivered at a health facility
2. Consent and privacy for vaginal examinations
3. Appropriate pain relief ^e^
4. The proportion who had adequate privacy during delivery using curtains, partitions or other measures
5. Overall rating of the quality of delivery visits as very good/excellent
6. The overall likelihood of recommending the health care provider to others for childbirth ^f^

###### Immediate postnatal care

1. The proportion who had a postnatal check-up before discharge
2. Immediate postnatal counselling index ^g^

###### Continuity of care

1. A continuity of care index calculated from the proportion who had;
  a. their first ANC visit during the first trimester,
  b. four or more ANC visits during the entire pregnancy,
  c. delivered at a health facility and
  d. received a postnatal check-up before discharge

*^a^ First ANC visit screening and treatment index included: measurements of blood pressure, weight, height, and mid-upper arm circumference (MUAC), as well as tests for HIV, syphilis, blood sugar, urine, and haemoglobin levels, iron and folic acid supplementation (IFAS) and tetanus injection across all countries and calcium supplementation in Ethiopia and South Africa*.

*^b^ First ANC visit counselling index included being counselled on danger signs during pregnancy, nutrition, physical exercise, birth plan and scheduling of follow-up visits*.

*^c^ Women were asked to rate the overall quality of their first ANC visit on a 5-point Likert scale from “very poor” to “excellent”*.

*^d^ Women who continued to take IFAS at least once during follow-up ANC*.

*^e^ Included requesting or was given or received pain relief during delivery*.

*^f^ Women were asked to rate whether they were likely to recommend their birth facility or provider to a family member or friend for childbirth on a 4-point Likert scale from “not at all likely” to “very likely”*.

*^g^ Immediate postnatal counselling index included being counselled on what the baby should eat, umbilical cord care, keeping the baby warm, returning for baby vaccinations, hand washing with soap, danger signs for the baby and danger signs for the mother before discharge after delivery*.

#### Risk factors for adverse delivery outcomes (mistreatment during childbirth and obstetric complications) among adolescents

As adolescents are at an increased risk of being mistreated during childbirth and poor pregnancy outcomes, we assessed the factors associated with these two outcomes. To measure mistreatment, we used eight questions to evaluate the experience of physical or verbal abuse. Four questions were used to assess physical abuse, and four to assess verbal abuse as recommended by Leslie et al. (21) (see S1 Table).

To examine obstetric complications, we analysed key adverse outcomes women may encounter during delivery and the immediate postpartum period. These outcomes included prolonged labour exceeding 12 hours, admission to the intensive care unit (ICU), obstetric fistula, the need for blood transfusions, and self-reported severe health complications (22).

Lastly, we assessed the factors associated with the adverse delivery outcomes. The assessed factors included country and region of residence, delivery facility characteristics (such as level and ownership), and sociodemographic variables (age, marital status, educational attainment, wealth index, health literacy, and employment status).

Additionally, we examined reproductive and obstetric factors, including gravidity, gestational age at the first ANC visit, pregnancy intentionality, and medical or obstetric history, alongside the quality of ANC and user experience.

### Statistical analysis

Our primary focus was to compare key measures between adolescents and adults. Descriptive analyses were stratified by the two age groups. We used chi-square tests and t-tests to assess any differences in the quality of care between adults and adolescents. A p-value of <0.05 was considered statistically significant. To assess the factors associated with obstetric complications and mistreatment during delivery, we used logistic regression for the subsample of adolescents only. These analyses combined adolescent girls from all three countries and included countries as fixed effects. We undertook bivariable and multivariable regression analyses. The results are presented as the crude and adjusted odds ratio (OR, with 95% CI). A p-value of <0.05 was considered statistically significant. All analyses were conducted using Stata version 17 (StataCorp, College Station, TX, USA).

### Ethical approval

The study protocol was reviewed and approved by the Institutional Review Boards (IRB) of the Harvard T.H. Chan School of Public Health (protocol #IRB22-0487), the Kenya Medical Research Institute (protocol number KEMRI/SERU/CGMR-C/4226), the Ethiopian Public Health Institute (protocol number EPHI-IRB-448-2022) and the University of KwaZulu-Natal (protocol number BREC/00004645/2022). Formal written informed consent was obtained from all adult study participants or emancipated minors or from formal guardians or next-of-kins for study participants under 18 years old per local regulations.

## RESULTS

### Sociodemographic factors

Table 1 summarises the demographic characteristics of 3,051 women included in the study. The number of adolescent girls ranged from 86 to 173 across the three countries, with a mean age of 18. In contrast, adult women had a mean age of 28 years. Among adolescents, 13% to 24% were aged 15–17, while 76% to 88% were aged 18–19. Marital status varied significantly: most adolescents in Kenya and South Africa were unmarried, whereas in Ethiopia, 95% were married. Educational attainment also showed notable differences. Nearly two-thirds of Ethiopian adolescents had either no education or only some primary education. Conversely, over half of Kenyan and South African adolescents completed primary education. Across all countries, a more significant proportion of adolescents reside in rural areas.

**Table 1.** Sociodemographic characteristics of pregnant adolescents and adult women enrolled in the Maternal and Newborn Health (MNH) eCohort longitudinal study in Ethiopia, Kenya and South Africa, 2023-2024

|  | Ethiopia |  | Kenya |  | South Africa |  | Total |  |  |
| --- | --- | --- | --- | --- | --- | --- | --- | --- | --- |
|  | Adolescent<br>(N=86) | Adults<br>(N=914) | Adolescents<br>(N=121) | Adults<br>(N=888) | Adolescents<br>(N=173) | Adults<br>(N=869) | Adolescents<br>(N=380) | Adults<br>(N=2,671) | Total<br>(N=3,051) |
| **Age, mean (SD)** | 18.2 (0.9) | 26.1 (4.5) | 18.0 (1.0) | 28.0 (5.7) | 18.0 (0.97) | 28.4 (5.71) | 18.1 (0.9) | 27.5 (5.4) | 26.3 (6.0) |
| **Marital status: “Married or partnered”, n (%)** | 82 (95.4) | 890 (97.5) | 57 (47.5) | 730 (82.5) | 2 (1.2) | 124 (14.3) | 141 (37.2) | 1744 (65.4) | 1885 (61.9) |
| **Education level, n (%)** |  |  |  |  |  |  |  |  |  |
| None or some primary education | 54 (62.8) | 330 (36.1) | 8 (6.6) | 62 (7.0) | 1 (0.6) | 10 (1.2) | 63 (16.6) | 402 (15.1) | 465 (15.3) |
| Completed primary education | 31 (36.0) | 364 (39.9) | 71 (58.7) | 280 (31.5) | 89 (51.5) | 206 (23.7) | 191 (50.3) | 850 (31.9) | 1041 (34.1) |
| Completed secondary or higher | 1 (1.2) | 219 (24.0) | 42 (34.7) | 546 (61.5) | 83 (48.0) | 652 (75.2) | 126 (33.2) | 1417 (53.1) | 1543 (50.6) |
| **Wealth index, n (%)** |  |  |  |  |  |  |  |  |  |
| Poorest | 32 (37.6) | 166 (18.5) | 48 (39.7) | 154 (17.5) | 40 (23.1) | 224 (25.8) | 120 (31.7) | 544 (20.6) | 664 (22.0) |
| Middle | 48 (56.5) | 166 (18.5) | 63 (52.1) | 545 (61.8) | 72 (41.6) | 338 (38.9) | 183 (48.3) | 1423 (53.8) | 1606 (53.1) |
| Richest | 5 (5.9) | 540 (60.3) | 10 (8.3) | 183 (20.8) | 61 (35.3) | 306 (35.3) | 76 (20.1) | 679 (25.7) | 755 (25.0) |
| **Region, n (%)** |  |  |  |  |  |  |  |  |  |
| Rural | 60 (69.8) | 448 (49.0) | 89 (73.6) | 418 (47.0) | 115 (66.5) | 400 (46.0) | 264 (69.5) | 1265 (47.4) | 1529 (50.1) |
| **Very good health literacy levels, n (%)** | 5 (5.8) | 261 (28.6) | 35 (28.9) | 416 (46.8) | 35 (20.2) | 310 (35.7) | 75 (19.7) | 987 (37.0) | 1062 (34.8) |
| **Employment status, n (%)** |  |  |  |  |  |  |  |  |  |
| Employed | 16 (18.6) | 371 (40.6) | 24 (20.2) | 522 (58.8) | 0 | 193 (22.2) | 40 (10.6) | 1086 (40.7) | 1126 (37.0) |
| **Social support, mean (SD)** | 3.3 (2.4) | 3.0 (2.2) | 3.3 (2.1) | 3.2 (2.2) | 2.5 (1.5) | 2.6 (1.8) | 3.0 (2.0) | 3.0 (2.1) | 3.0 (2.1) |
| **Gravidity mean (SD)** | 1.2 (0.5) | 2.6 (1.4) | 1.2 (0.6) | 2.7 (1.5) | 1.1 (0.3) | 2.5 (1.3) | 1.2 (0.5) | 2.6 (1.4) | 2.4 (1.4) |
| **Gestational age at first ANC visit, n (%)** |  |  |  |  |  |  |  |  |  |
| 1st trimester 0-12 weeks | 18 (21.4) | 286 (31.9) | 9 (7.4) | 147 (16.6) | 59 (34.3) | 366 (42.2) | 86 (22.8) | 799 (30.1) | 885 (29.2) |
| 2nd trimester 13-26 weeks | 60 (71.4) | 555 (61.9) | 74 (61.2) | 564 (63.5) | 89 (51.7) | 418 (48.2) | 223 (59.2) | 1537 (58.0) | 1760 (58.1) |
|  | Adolescent<br>(N=86) | Adults<br>(N=914) | Adolescents<br>(N=121) | Adults<br>(N=888) | Adolescents<br>(N =173) | Adults<br>(N=869) | Adolescents<br>(N=380) | Adults<br>(N=2,671) | Total<br>(N=3,051) |
| 3rd trimester 27-42 weeks | 6 (7.1) | 55 (6.1) | 38 (31.4) | 177 (19.9) | 24 (14.0) | 84 (9.7) | 68 (18.0) | 316 (11.9) | 384 (12.7) |
| **Pregnancy was intended, n (%)** | 61 (70.9) | 667 (73.1) | 48 (40.0) | 567 (64.7) | 9 (5.2) | 172 (19.8) | 118 (31.2) | 1405 (52.9) | 1523 (52.9) |
| **Current medical/obstetric history, n (%)** |  |  |  |  |  |  |  |  |  |
| Underweight (MUAC < 23 cm or BMI <18.5) | 33 (38.4) | 239 (26.2) | 9 (7.4) | 37 (4.2) | 5 (2.9) | 12 (1.4) | 47 (12.4) | 288 (10.8) | 335 (11.0) |
| Experienced at least one pregnancy danger sign | 21 (24.4) | 221 (24.2) | 25 (20.7) | 208 (23.4) | 82 (47.7) | 400 (46.4) | 128 (33.8) | 829 (31.1) | 957 (31.4) |
| Experienced ANC depression (PHQ9>5) | 16 (18.6) | 231 (25.3) | 21 (17.4) | 179 (20.2) | 34 (20.0) | 116 (13.4) | 71 (18.8) | 526 (19.7) | 597 (19.6) |
| Rates own health as poor or fair | 10 (11.6) | 160 (17.5) | 7 (5.8) | 80 (9.0) | 18 (10.4) | 64 (7.4) | 35 (9.2) | 304 (11.4) | 339 (11.1) |
| Engaged in risky health behaviour (substance use) | 15 (17.4) | 224 (24.6) | 3 (2.5) | 14 (1.6) | 11 (6.4) | 82 (9.4) | 29 (7.6) | 320 (12.0) | 349 (11.5) |
| **Experienced Intimate Partner Violence**<br>**(emotional/physical), n (%)** | 7 (8.1) | 58 (6.3) | 10 (8.3) | 60 (6.7) | 18 (10.4) | 48 (5.5) | 35 (9.2) | 166 (6.2) | 201 (6.6) |

\*SD: Standard deviation; ANC: Antenatal care; MUAC: Mid-upper arm circumference; BMI: Body Mass Index; PHQ9: Patient Health Questionnaire-9

Most adolescents initiated ANC in the second trimester. The average number of pregnancies was one among adolescents and three among adults across all countries. In Ethiopia, over 70% of adolescents reported that their pregnancies were intentional, compared to only 5.2% and 40% in South Africa and Kenya. Additionally, 21% to 48% of adolescents reported experiencing at least one danger sign during pregnancy. Reports of intimate partner violence or abuse were 7%, ranging from 8% to 10% across both groups and all countries (Table 1).

### Quality and continuity of maternal health care

Table 2 shows the quality of antenatal, delivery, and postnatal care, while Fig 1 illustrates the proportion of women receiving care along the continuum of maternal health care across the three countries.

**Table 2.** Quality of maternal health care received by adolescents compared to adult women among women enrolled in the MNH eCohort longitudinal study in Ethiopia, Kenya and South Africa, 2023-2024

|  | Ethiopia |  |  | Kenya |  |  | South Africa |  |  | Total |  |  |
| --- | --- | --- | --- | --- | --- | --- | --- | --- | --- | --- | --- | --- |
|  | Adolescents | Adults | P-value | Adolescents | Adults | P-value | Adolescents | Adults | P-value | Adolescents | Adults | P-value |
|  | N=86 | N=914 |  | N=121 | N=888 |  | N=173 | N=876 |  | N=380 | N=2,671 |  |
| **ANTENATAL CARE** |  |  |  |  |  |  |  |  |  |  |  |  |
| **First ANC visit** |  |  |  |  |  |  |  |  |  |  |  |  |
| First ANC completeness index: screening and treatments (mean (SD)) | 49.4 (14.9) | 50.9 (15.5) | 0.367 | 70.9 (15.1) | 69.3 (14.5) | 0.270 | 85.4 (7.9) | 84.6 (8.7) | 0.644 | 72.6 (18.6) | 68.0 (19.2) | <0.001 |
| First ANC Completeness index: counselling (mean (SD)) | 31.3 (18.9) | 33.7 (20.7) | 0.280 | 50.3 (30.0) | 60.6 (29.6) | <0.001 | 56.7 (27.6) | 57.8 (27.8) | 0.865 | 48.9 (28.5) | 50.5 (28.9) | 0.310 |
| **Follow-up (FU) ANC visits** |  |  |  |  |  |  |  |  |  |  |  |  |
| Number of additional ANC visits (mean, SD) | 2.6 (1.7) | 3.1 (1.9) | 0.033 | 1.8 (1.5) | 2.1 (1.8) | 0.042 | 1.8 (2.0) | 2.2 (2.3) | 0.406 | 2.0 (1.8) | 2.5 (2.1) | <0.001 |
| Total number of ANC visits (mean, SD) | 3.9 (1.8) | 4.7 (2.2) | 0.002 | 2.9 (1.6) | 3.3 (1.9) | 0.032 | 2.8 (2.0) | 3.2 (2.3) | 0.406 | 3.1 (1.9) | 3.8 (2.3) | <0.001 |
| At least one blood test during FU ANC (n (%)) | 34 (39.5) | 446 (48.8) | 0.100 | 43 (35.5) | 315 (35.5) | 0.989 | 93 (53.8) | 523 (60.2) | 0.116 | 170 (44.7) | 1,284 (48.1) | 0.223 |
| At least one urine test during FU ANC (n (%)) | 21 (24.4) | 308 (33.7) | 0.080 | 21 (17.4) | 234 (26.4) | 0.033 | 99 (57.2) | 578 (66.5) | 0.019 | 141 (37.1) | 1,120 (41.9) | 0.074 |
| Received food supplementation at last once (n (%)) | 3 (3.5) | 42 (4.6) | 0.636 | 0 | 18 (2.0) | 0.114 | 6 (3.5) | 22 (2.5) | 0.487 | 9 (2.4) | 82 (3.1) | 0.452 |
| Ultrasound done at one point (baseline and follow up) (n (%)) | 73 (84.9) | 803 (87.9) | 0.424 | 53 (43.8) | 478 (53.8) | 0.038 | 64 (37.0) | 418 (48.1) | 0.007 | 190 (50.0) | 1,699 (63.6) | <0.001 |
| Continues to take Iron and folic acid (adherence at least once during FU) (n (%)) | 59 (68.6) | 676 (74.0) | 0.282 | 64 (52.9) | 580 (65.3) | 0.008 | 101 (58.4) | 598 (68.8) | 0.008 | 224 (59.0) | 1,854 (69.4) | <0.001 |
| Counselling on postpartum family planning at least once during FU ANC (n (%)) | 0 | 40 (4.4) | 0.048 | 11 (9.1) | 153 (17.2) | 0.023 | 67 (38.7) | 396 (45.6) | 0.098 | 78 (20.5) | 589 (22.1) | 0.501 |
| **DELIVERY CARE (among women who gave birth in facilities)** | **N=64** | **N=743** |  | **N=95** | **N=733** |  | **N=138** | **N=665** |  | **N=297** | **N=2,141** |  |
| **Consent and privacy for vaginal exams (n (%))** |  |  |  |  |  |  |  |  |  |  |  |  |
| Vaginal examination | 29 (45.3) | 316 (42.5) | 0.666 | 71 (74.7) | 491 (67.0) | 0.128 | 109 (79.0) | 515 (77.4) | 0.692 | 209 (70.4) | 1,322 (61.8) | 0.004 |
| Vaginal examination with permission | 12 (18.8) | 153 (20.6) | 0.726 | 54 (56.8) | 383 (52.3) | 0.399 | 78 (56.5) | 440 (66.2) | 0.031 | 144 (48.5) | 976 (45.6) | 0.348 |
| Privacy during vaginal examination | 20 (31.3) | 234 (31.5) | 0.968 | 66 (69.5) | 464 (63.3) | 0.238 | 102 (73.9) | 490 (73.7) | 0.956 | 118 (63.3) | 1,188 (55.5) | 0.011 |
| **Appropriate pain relief** |  |  |  |  |  |  |  |  |  |  |  |  |
| Requested for pain relief | 8 (12.5) | 159 (21.4) | 0.092 | 21 (22.1) | 228 (31.1) | 0.072 | 27 (19.6) | 152 (22.9) | 0.398 | 56 (18.9) | 539 (25.2) | 0.017 |
|  | Adolescents | Adults | P-value | Adolescents | Adults | P-value | Adolescents | Adults | P-value | Adolescents | Adults | P-value |
| Offered/received pain relief | 13 (20.3) | 254 (34.2) | 0.024 | 43 (45.3) | 511 (69.7) | <0.001 | 121 (87.7) | 594 (89.3) | 0.574 | 177 (59.6) | 1,359 (63.5) | 0.194 |
| Privacy during delivery (n (%)) | 35 (57.8) | 445 (61.9) | 0.492 | 87 (91.6) | 677 (92.4) | 0.593 | 105 (77.8) | 561 (86.3) | 0.012 | 227 (76.4) | 1,682 (78.6) | 0.672 |
| Mistreatment during delivery (physical/mental) (n (%)) | 23 (39.0) | 182 (26.0) | 0.030 | 14 (14.7) | 79 (10.8) | 0.497 | 43 (31.4) | 138 (21.1) | 0.009 | 80 (26.9) | 399 (18.6) | 0.003 |
| **IMMEDIATE POSTPARTUM CARE (among women who gave birth in facilities)** | **N=59** | **N=698** |  | **N=94** | **N=728** |  | **N=138** | **N=658** |  | **N=291** | **N=2,084** |  |
| Postnatal check-up before discharge | 21 (35.6) | 412 (59.0) | <0.001 | 87 (92.6) | 672 (92.3) | 0.933 | 119 (86.2) | 602 (91.5) | 0.055 | 227 (78.0) | 1,686 (80.9) | 0.242 |
| **Immediate postnatal counselling** | **N=61** | **N=704** |  | **N=92** | **N=719** |  | **N=137** | **N=649** |  | **N=290** | **N=2,070** |  |
| What the baby should eat | 17 (27.9) | 269 (38.3) | 0.108 | 74 (80.4) | 631 (87.9) | 0.045 | 92 (67.2) | 495 (76.4) | 0.024 | 183 (63.1) | 1,395 (67.4) | 0.143 |
| Umbilical cord care | 16 (26.2) | 250 (35.5) | 0.144 | 79 (85.9) | 675 (94.0) | 0.004 | 100 (73.0) | 488 (75.4) | 0.673 | 195 (67.2) | 1,413 (68.3) | 0.719 |
| Keep baby warm | 7 (11.5) | 208 (29.6) | 0.003 | 72 (78.3) | 626 (87.3) | 0.018 | 84 (61.3) | 441 (68.0) | 0.550 | 163 (56.2) | 1,275 (61.7) | 0.075 |
| Return for baby vaccinations | 35 (57.4) | 449 (63.9) | 0.313 | 85 (92.4) | 691 (96.2) | 0.083 | 123 (89.8) | 599 (92.3) | 0.328 | 243 (83.8) | 1,739 (84.0) | 0.925 |
| Hand washing with soap | 12 (19.7) | 181 (25.8) | 0.292 | 73 (79.4) | 589 (82.0) | 0.530 | 93 (67.9) | 464 (71.5) | 0.398 | 178 (61.4) | 1,234 (59.4) | 0.572 |
| Danger signs for the baby | 13 (21.3) | 185 (26.4) | 0.389 | 59 (64.1) | 524 (73.0) | 0.075 | 77 (56.2) | 419 (64.7) | 0.062 | 149 (51.4) | 1,128 (54.6) | 0.311 |
| Danger signs for the mother | 14 (23.0) | 203 (28.9) | 0.325 | 55 (59.8) | 510 (70.9) | 0.028 | 77 (56.2) | 408 (63.2) | 0.128 | 146 (50.3) | 1,121 (54.2) | 0.211 |
| Immediate postpartum counselling completeness index (mean (SD)) | 26.7 (28.4) | 35.5 (32.5) | 0.041 | 77.2 (29.01) | 84.5 (23.5) | 0.006 | 67.4 (29.9) | 73.1 (30.1) | 0.043 | 61.9 (34.7) | 64.2 (35.8) | 0.301 |
| **USER EXPERIENCE** |  |  |  |  |  |  |  |  |  |  |  |  |
| **First ANC visit** | **N=86** | **N=914** |  | **N=121** | **N=888** |  | **N=173** | **N=869** |  | **N=380** | **N=2,671** |  |
| Overall rating of the quality of first ANC visits as very good/excellent | 49 (57.0) | 487 (53.3) | 0.511 | 72 (59.5) | 560 (63.1) | 0.448 | 84 (48.6) | 485 (55.8) | 0.080 | 205 (54.0) | 1,532 (57.4) | 0.209 |
| **Delivery care** | **N=61** | **N=715** |  | **N=94** | **N=727** |  | **N=138** | **N=665** |  | **N=293** | **N=2,107** |  |
| Overall rating of the quality of delivery care as very good/excellent | 31 (50.8) | 308 (43.1) | 0.242 | 43 (45.7) | 380 (52.3) | 0.234 | 64 (46.4) | 346 (52.0) | 0.227 | 138 (47.1) | 1,034 (49.1) | 0.526 |
| Very likely to recommend the provider for childbirth | 41 (68.3) | 391 (55.2) | 0.048 | 55 (58.5) | 458 (63.0) | 0.398 | 78 (56.5) | 393 (59.6) | 0.511 | 174 (59.6) | 1,242 (59.3) | 0.914 |

**Fig 1.**
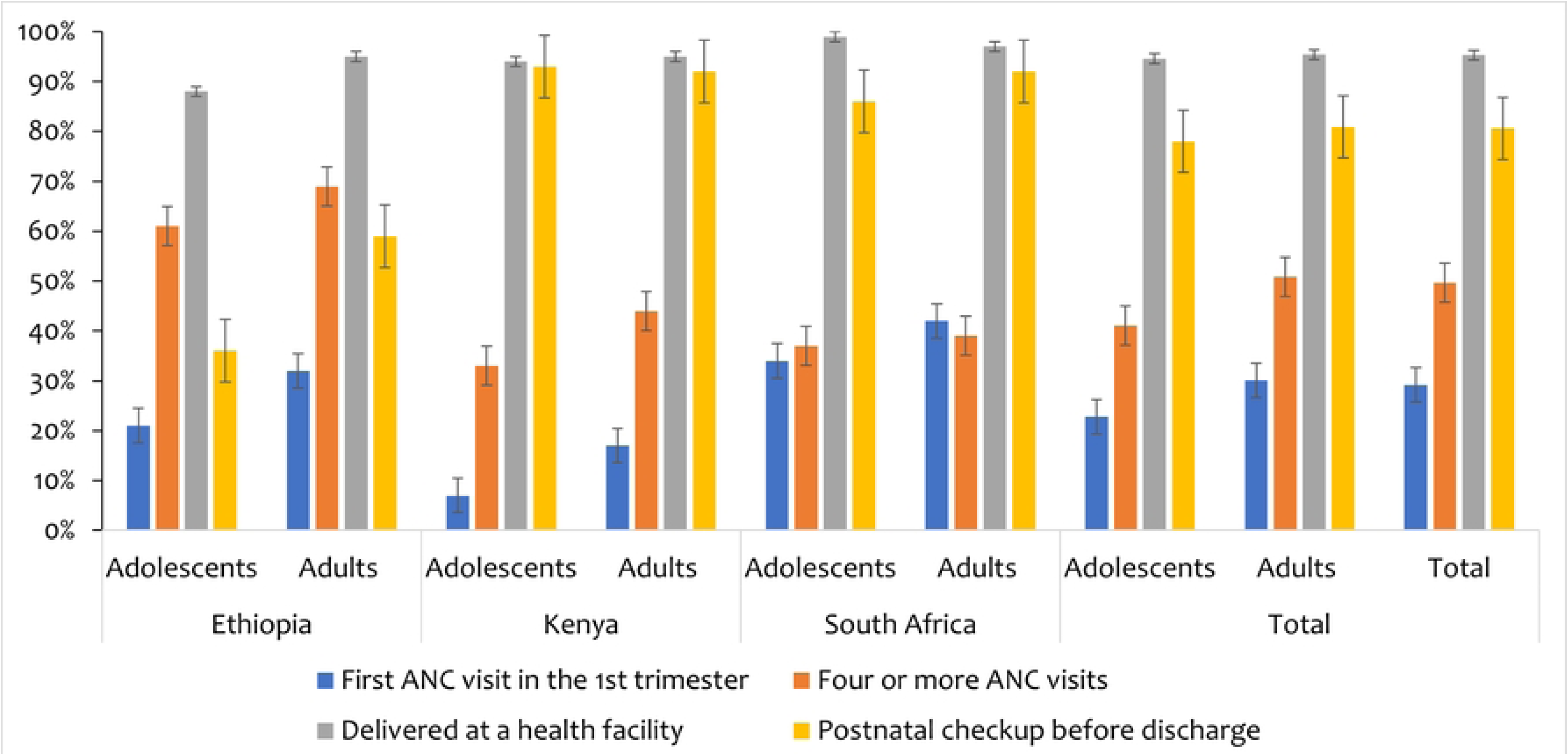
Continuity of maternal health care between adolescents and adults enrolled in the MNH eCohort longitudinal study in Ethiopia, Kenya and South Africa, 2023-2024

#### Antenatal care

Less than one-third of women attended their first ANC visit during the first trimester, with significantly higher rates among adults (30%) compared to adolescents (22%, p=0.003). This disparity was particularly evident in Kenya (17% vs. 7%, p=0.009) and South Africa (42% vs. 34%, p=0.050), where adult women were more likely than adolescents to initiate ANC early (Fig 1).

The overall completeness of screenings and treatments during the first ANC visit was 69%, with adolescents receiving significantly more comprehensive care than adults. Completeness levels varied between 50% and 85% across the three countries, with no substantial differences between age groups within individual countries. However, the completeness of ANC counselling was lower, averaging 50% and ranging from 31% to 61% across all settings. Notably, in Kenya, adults had significantly higher counselling rates than adolescents (61% vs. 50%, p<0.001). With these variations in service delivery, just over half of the women rated the quality of their first ANC visit as very good or excellent, with no significant differences between adolescents and adults across all countries (Table 2).

On average, women attended two follow-up (FU) ANC visits, with adult women significantly more likely than adolescents to attend additional visits (3 vs. 2, p<0.001), particularly in Ethiopia and Kenya. Less than half of all women received additional blood (48%) or urine tests (41%) during FU visits. However, adolescents were significantly less likely than adults to receive urine tests in Kenya (17% vs. 26%, p=0.033) and South Africa (57% vs. 67%, p=0.019) (Table 2).

Over two-thirds of all women received an ultrasound during ANC, with adult women significantly more likely than adolescents to undergo the procedure (64% vs. 50%, p<0.001). While ultrasound coverage was high in Ethiopia, where over 80% of both groups received at least one scan, adolescents were significantly less likely than adults to receive ultrasounds in Kenya (44% vs. 54%, p=0.038) and South Africa (37% vs. 48%, p=0.007). In contrast, food supplementation during ANC visits was rare, with only 3% of women reporting to have received it (Table 2).

Adherence to iron-folic acid supplementation (IFAS) during FU ANC visits was 68% overall, with significantly higher adherence among adult women compared to adolescents (69% vs. 59%, p<0.001). This disparity was particularly pronounced in Kenya (65% vs. 53%, p=0.008) and South Africa (69% vs. 58%, p=0.008). However, adherence rates were relatively high in Ethiopia, where more than two-thirds of both groups continued IFAS use. Regarding postpartum family planning, only 22% of women received counselling at least once during FU ANC visits, with significantly lower rates among adolescents compared to adults in Ethiopia and Kenya (Table 2).

The total number of ANC visits averaged four, ranging from three to five across all age groups and countries. However, adult women attended significantly more ANC visits than adolescents, particularly in Kenya and Ethiopia. Nearly half of all women attended more than four ANC visits, with adult women significantly more likely than adolescents to do so (51% vs. 41%, p<0.001), with notable differences observed in Kenya (Table 2).

#### Delivery care

Over 95% of women delivered in a health facility, with facility-based delivery rates ranging from 88% to 99% across both age groups and the three countries. However, significant differences were observed in Ethiopia, where adult women were significantly more likely than adolescents to deliver in a health facility (95% vs. 88%, p=0.016) (Fig 1).

More than 60% of women reported undergoing vaginal examinations, with adolescents experiencing these procedures at a higher rate than adults (70% vs. 62%, p=0.004). Despite this, informed consent for vaginal examinations remained relatively low across all three countries, with only 45% of women reporting that they were asked for permission before the procedure. Significant differences were observed in South Africa, where adolescents were less likely than adults to report being asked for consent (p=0.031). Additionally, just over half of the women indicated that privacy was maintained during vaginal examinations, with adolescents reporting significantly higher rates of privacy compared to adult women (63% vs. 56%, p=0.011). In contrast, privacy during delivery was reported by over 78% of women. However, significant differences were noted in South Africa, where adult women were more likely than adolescents to report having privacy (86% vs. 78%, p=0.012) (Table 2).

Regarding pain relief during delivery, only 24% of women requested it, with a lower proportion of adolescents making such requests in South Africa. Nevertheless, over 60% of women reported either receiving or being offered pain relief. Despite this, significant disparities existed, with adolescents having lower access to pain relief than adults in Ethiopia (20% vs. 34%, p=0.024) and Kenya (45% vs. 70%, p<0.001). In contrast, both groups in South Africa reported high levels of pain relief, with no significant difference between adolescents (88%) and adults (89%, p=0.574) (Table 2).

Less than half of the women (49%) rated the quality of delivery care as very good or excellent, with no significant differences between adolescents and adults. However, 59% of women indicated a high likelihood of recommending their childbirth provider to others. In Ethiopia, adolescents were significantly more likely than adults to recommend their provider (68% vs. 55%, p=0.048) (Table 2).

#### Immediate postpartum care

Among women who delivered in a health facility, 81% reported receiving a postnatal check-up before discharge. However, rates were significantly lower in Ethiopia, where just over one-third of adolescents and more than half of adults received a postnatal check-up (Table 2 and Fig 1).

On average, women received 64% of the recommended immediate postnatal counselling messages, with adult women receiving significantly more counselling than adolescents across all three countries. Adults were more likely to be counselled on key postnatal topics like infant feeding, umbilical cord care, keeping the baby warm, and recognising maternal danger signs, particularly in Kenya (Table 2).

The overall continuity of care index—defined as attending the first ANC visit in the first trimester, completing at least four ANC visits, delivering in a health facility, and receiving a postnatal check-up before discharge—was 58 across all women, with adults having a significantly higher score than adolescents (59 vs. 53, p<0.001). This trend was consistent across all three countries, with significant differences observed in Ethiopia (60 vs. 48, p<0.001) and Kenya (56 vs. 49, p=0.003), while no significant difference was found in South Africa (61 vs. 58, p=0.313) (Fig 1).

### Experience of adverse delivery outcomes

Table 2 presents the prevalence of mistreatment and obstetric complications among adolescent girls and adult women. Overall, 20% of women reported experiencing mistreatment, including physical or verbal abuse during delivery, with significantly higher rates among adolescents compared to adults (27% vs. 19%, p=0.003). Adolescents reported higher levels of mistreatment than adults in Ethiopia (39% vs. 26%, p=0.030) and South Africa (31% vs. 21%, p=0.009) (Table 3).

**Table 3.** Prevalence of adverse delivery outcomes between adolescents and adults enrolled in the MNH eCohort longitudinal study in Ethiopia, Kenya and South Africa, 2023-2024

|  | Ethiopia |  |  | Kenya |  |  | South Africa |  |  | Total |  |  |  |
| --- | --- | --- | --- | --- | --- | --- | --- | --- | --- | --- | --- | --- | --- |
|  | Adolescents<br>N= 64 | Adults<br>N=743 | P-value | Adolescents<br>N=95 | Adults<br>N=733 | P-value | Adolescents<br>N = 138 | Adults<br>N=665 | P-value | Adolescents<br>N = 297 | Adults<br>N=2,141 | P-value | Total<br>N=2,438 |
|  | n (%) | n (%) |  | n (%) | n (%) |  | n (%) | n (%) |  | n (%) | n (%) |  | n (%) |
| Mistreatment during delivery<br>(physical/mental) | 23 (39.0) | 182 (26.0) | 0.030 | 14 (14.7) | 79 (10.8) | 0.497 | 43 (31.4) | 138 (21.1) | 0.009 | 80 (26.9) | 399<br>(18.6) | 0.003 | 479<br>(19.7) |
| Caesarian section | 7 (10.9) | 158 (21.3) | 0.135 | 16 (16.8) | 222 (30.3) | 0.006 | 42 (30.4) | 246 (37.0) | 0.238 | 65 (21.9) | 526<br>(29.2) | 0.026 | 691<br>(28.2) |
| Episiotomy | 29 (45.3) | 170 (22.9) | <0.001 | 44 (46.3) | 160 (21.8) | <0.001 | 75 (54.4) | 189 (28.4) | <0.001 | 148 (49.8) | 519<br>(24.2) | <0.001 | 667<br>(27.4) |
| **Other obstetric complications** |  |  |  |  |  |  |  |  |  |  |  |  |  |
| Prolonged labor (over 12 hours) | 14 (23.4) | 130 (17.5) | 0.310 | 18 (18.0) | 132 (18.0) | 0.823 | 31 (22.5) | 80 (12.0) | 0.001 | 64 (21.6) | 342<br>(16.0) | 0.054 | 406<br>(16.7) |
| Admitted to ICU | 3 (4.7) | 9 (1.2) | 0.076 | 1 (1.1) | 12 (1.6) | 0.666 | 5 (3.6) | 13 (2.0) | 0.228 | 9 (3.0) | 34 (1.6) | 0.201 | 43 (1.8) |
| Blood transfusion | 1 (1.6) | 12 (1.6) | 0.579 | 2 (2.1) | 26 (3.6) | 0.464 | 15 (10.9) | 37 (5.6) | 0.033 | 18 (6.1) | 75<br>(3.5) | 0.093 | 93 (3.8) |
| Obstetric fistula | 1 (1.6) | 8 (1.1) | 0.854 | 1 (1.1) | 8 (1.1) | 0.973 | 5 (3.6) | 14 (2.1) | 0.307 | 7 (2.4) | 30 (1.4) | 0.355 | 37 (1.5) |
| Self-reported severe health problems | 3 (4.7) | 89 (12.0) | 0.030 | 1 (1.1) | 49 (6.7) | 0.034 | 9 (6.5) | 51 (7.7) | 0.329 | 13 (4.4) | 189<br>(8.8) | 0.015 | 202<br>(8.3) |
| At least 1 other obstetric<br>complication <sup>a</sup> | 18 (28.1) | 202 (27.2) | 0.872 | 20 (21.1) | 191 (26.1) | 0.292 | 54 (39.1) | 162 (24.3) | <0.001 | 92 (31.0) | 255<br>(25.9) | 0.065 | 647<br>(26.5) |
| Peripartum depression (PHQ2>3) | 2 (3.1) | 76 (10.2) | 0.174 | 3 (3.2) | 39 (5.3) | 0.664 | 11 (8.0) | 37 (5.6) | 0.275 | 16 (5.4) | 152<br>(7.1) | 0.409 | 46 (1.9) |

<sup>a</sup> Included the reporting of at least one of the other obstetric complications

Approximately 28% of women underwent a caesarean section, while 27% experienced an episiotomy. Caesarean section rates were significantly higher among adult women (29% vs. 22%, p=0.026), whereas episiotomies were substantially more common among adolescents (50% vs. 24%, p<0.001). Country-specific differences were observed, with caesarean sections being more frequent among adults in Kenya, while episiotomy rates were significantly higher among adolescents across all three countries (Table 3).

In addition, 27% of women reported experiencing at least one obstetric complication (other than c-sections or episiotomies) during delivery. In South Africa, adolescents had significantly higher rates of obstetric complications than adults (39% vs. 24%, p<0.001). Prolonged labour (lasting more than 12 hours) was the most frequently reported complication (17%), while admission to the ICU was the least common (2%). Adolescents in South Africa were significantly more likely than adults to experience prolonged labour (23% vs. 12%, p=0.001) and require blood transfusions (11% vs. 6%, p=0.033). However, in Ethiopia and Kenya, adolescents reported significantly lower rates of severe health complications compared to adults (Table 3).

#### Factors associated with mistreatment during delivery among adolescents from Ethiopia, Kenya and South Africa

The bivariable analysis revealed various factors associated with mistreatment during delivery among adolescents. Rural residence (OR: 3.15, 95% CI: 1.60–6.19), antenatal depression (OR: 2.28, 95% CI: 1.20–4.31), engaging in risky behaviour like substance use (OR: 3.53, 95% CI: 1.46–8.54), and intimate partner violence (OR: 5.36, 95% CI: 2.15–13.3) were associated with higher odds of mistreatment. Conversely, being in Kenya (OR: 0.27, 95% CI: 0.13–0.59), having good health literacy (OR: 0.41, 95% CI: 0.20–0.86), and having privacy during delivery (OR: 0.24, 95% CI: 0.14–0.44) was linked to lower odds of mistreatment (Table 4).

**Table 4:** Results of logistic regression models for the factors associated with experiencing any mistreatment during delivery among adolescents enrolled in the MNH eCohort longitudinal study in Ethiopia, Kenya and South Africa, 2023-2024

|  | N=290 |  |  | N=280 |  |  |
| --- | --- | --- | --- | --- | --- | --- |
| **Factors** | **Crude OR** | **95% CI** | **p-value** | **aOR** | **95% CI** | **p-value** |
| **Country** |  |  |  |  |  |  |
| Ethiopia | Ref |  |  |  |  |  |
| Kenya | **0.27** | **0.13 – 0.59** | **0.001** | **0.19** | **0.05 – 0.75** | **0.018** |
| South Africa | 0.72 | 0.38 – 1.35 | 0.303 | 0.36 | 0.08 – 1.75 | 0.207 |
| **Region** |  |  |  |  |  |  |
| Urban | Ref |  |  |  |  |  |
| Rural | **3.15** | **1.60 – 6.19** | **0.001** | **2.61** | **1.11 – 6.13** | **0.028** |
| **Facility ownership** |  |  |  |  |  |  |
| FBO/NGO/Private | Ref |  |  |  |  |  |
| Government | 1.79 | 0.77 – 4.16 | 0.175 | **4.33** | **1.44 – 13.1** | **0.009** |
| **Health literacy levels** |  |  |  |  |  |  |
| Good/Fair/Poor | Ref |  |  |  |  |  |
| Excellent/Very good | **0.41** | **0.20 – 0.86** | **0.018** | 0.65 | 0.27 – 1.57 | 0.339 |
| **Experienced ANC depression (PHQ9>5)** |  |  |  |  |  |  |
| No | Ref |  |  |  |  |  |
| Yes | **2.28** | **1.20 – 4.31** | **0.012** | 2.05 | 0.95 – 4.44 | 0.067 |
| **Engaged in risky health behaviour (substance use)** |  |  |  |  |  |  |
| No | Ref |  |  |  |  |  |
| Yes | **3.53** | **1.46 – 8.54** | **0.005** | 2.36 | 0.79 – 7.05 | 0.125 |
| **Experienced Intimate Partner Violence (emotional/physical)** |  |  |  |  |  |  |
| Factors | Crude OR | 95% CI | p-value | aOR | 95% CI | p-value |
| No | Ref |  |  |  |  |  |
| Yes | **5.36** | **2.15 – 13.3** | **<0.001** | **3.31** | **1.16 – 9.39** | **0.025** |
| **Privacy during delivery** |  |  |  |  |  |  |
| No | Ref |  |  |  |  |  |
| Yes | **0.24** | **0.14 – 0.44** | **<0.001** | **0.44** | **0.21 – 0.92** | **0.030** |
| **User experience** |  |  |  |  |  |  |
| **Overall rating of the quality of delivery care** |  |  |  |  |  |  |
| Poor/fair/Good | Ref |  |  |  |  |  |
| Excellent/Very good | **0.38** | **0.22 – 0.65** | **<0.001** | **0.37** | **0.18 – 0.76** | **0.007** |

From the multivariable analysis, adolescents from Kenya compared to those from Ethiopia (aOR: 0.19, 95% CI: 0.05–0.75), those who had privacy during delivery (aOR: 0.44, 95% CI: 0.21–0.92) and high-quality delivery care ratings (aOR: 0.38, 95% CI: 0.18–0.79) remained less likely to experience mistreatment. However, mistreatment was more common among adolescents from rural areas (aOR: 2.50, 95% CI: 1.07–5.84), those delivering in government-operated facilities (aOR: 4.40, 95% CI: 1.43–13.5), and those who experienced intimate partner violence (aOR: 3.49, 95% CI: 1.24–9.82). (Table 4).

#### Factors associated with obstetric complications among adolescents from Ethiopia, Kenya and South Africa

The bivariable analysis identified several factors associated with the odds of experiencing obstetric complications among adolescents. Higher odds of obstetric complications were associated with rural residence (OR: 1.91, 95% CI: 1.10–3.30), older age (18–19 years vs. 15– 17 years, OR: 1.54, 95% CI: 1.02–2.33), presence of at least one pregnancy danger sign (OR: 1.76, 95% CI: 1.09–2.84), and antenatal depression (OR: 1.75, 95% CI: 1.00–3.07). Conversely, delivery in a primary healthcare facility was associated with lower odds of complications (OR: 0.48, 95% CI: 0.28–0.83) (Table 5).

**Table 5:** Results of logistic regression models for the factors associated with experiencing at least one obstetric complication among adolescents enrolled in the MNH eCohort longitudinal study in Ethiopia, Kenya and South Africa, 2023-2024.

|  | N=380 |  |  | N=284 |  |  |
| --- | --- | --- | --- | --- | --- | --- |
| **Factors** | **Crude OR** | **95% CI** | **p-value** | **aOR** | **95% CI** | **p-value** |
| **Place of residence** |  |  |  |  |  |  |
| Urban | Ref |  |  |  |  |  |
| Rural | **1.91** | **1.10 – 3.30** | **0.022** | **2.25** | **1.09 – 4.63** | **0.028** |
| **Delivery Facility Characteristics** |  |  |  |  |  |  |
| **Facility level** |  |  |  |  |  |  |
| Secondary | Ref |  |  |  |  |  |
| Primary | **0.48** | **0.28 – 0.83** | **0.009** | 0.60 | 1.09 – 4.63 | 0.188 |
| **Age in years** |  |  |  |  |  |  |
| 15-17 | Ref |  |  |  |  |  |
| 18-19 | **1.54** | **1.02 – 2.33** | **0.040** | 1.26 | 0.70 – 2.28 | 0.445 |
| **Current medical/obstetric history** |  |  |  |  |  |  |
| Experienced at least one pregnancy danger sign |  |  |  |  |  |  |
| No | Ref |  |  |  |  |  |
| Yes | 1.76 | 1.09 – 2.84 | 0.021 | 1.12 | 0.61 – 2.04 | 0.711 |
| Experienced prenatal depression (PHQ9>5) |  |  |  |  |  |  |
| No | Ref |  |  |  |  |  |
| Yes | 1.75 | 1.00 – 3.07 | 0.049 | **2.43** | **1.19 – 5.00** | **0.015** |
| **Visited another facility before delivery facility** |  |  |  |  |  |  |
| No | Ref |  |  |  |  |  |
| Yes | 1.52 | 0.86 – 2.68 | 0.148 | **1.96** | **1.02 – 3.80** | **0.045** |

In the multivariable analysis, rural residence (aOR: 2.34, 95% CI: 1.13–4.84) and antenatal depression (aOR: 2.39, 95% CI: 1.17–4.89) remained significant predictors of obstetric complications. Additionally, adolescents who visited another health facility during labour had nearly twice the odds of reporting obstetric complications (aOR: 1.99, 95% CI: 1.03– 3.86) (Table 5).

## DISCUSSION

This study described the quality, continuity, and experiences of maternal healthcare among 380 pregnant adolescents compared to 2,671 pregnant adult women in Ethiopia, Kenya, and South Africa. We also investigated the factors associated with adverse delivery outcomes. The findings highlight critical gaps in care, with less than one-third of women initiating ANC in the first trimester and adults significantly more likely to do so than adolescents. While the completeness of the first ANC services was 69%, counselling rates were notably low at 50%. Moreover, follow-up ANC visits remained suboptimal, with adolescents receiving limited access to essential maternal health interventions such as ultrasounds, iron-folic acid supplementation, and postpartum family planning counselling. These disparities extended to delivery and postnatal care, where adolescents reported higher mistreatment, lower privacy, and reduced access to critical follow-up services. Additionally, rural residence, antenatal depression, and intimate partner violence significantly increased the likelihood of mistreatment and obstetric complications. Overall, quality of ANC, delivery and PNC, was often lower for adolescents compared to older women. Yet, perceived quality and satisfaction was similar, likely due to lower expectations from adolescents. Adolescents were also more likely to experience mistreatment during labor and delivery and more likely to receive an episiotomy, a potentially harmful practice.

Lower ANC utilisation, especially among adolescents, may be attributed to multiple factors, including stigma, lack of knowledge, transportation issues and financial constraints (16, 23, 24). Early ANC initiation is crucial for improving maternal outcomes, yet many adolescents delay seeking care due to fear of judgment from healthcare providers (17). In addition, most adolescent pregnancies were unplanned and therefore may have been detected late. Strengthening adolescent-friendly services, including community outreach programs and tailored health education, may improve early ANC uptake and continuity of care.

The results on facility delivery demonstrate encouraging trends, with over 80% of women delivering in health facilities across all three countries, higher than the rates reported across other countries (25). However, significant disparities persist in Ethiopia, where adult women were more likely to deliver in a facility compared to adolescents. These disparities suggest that adolescents may still face cultural or logistical barriers, such as fear of healthcare providers, financial constraints, or preference for traditional birth settings (16, 26, 27). Existing literature corroborates these findings, with studies in sub-Saharan Africa reporting that adolescent mothers are less likely to access facility-based delivery due to stigma and lack of support systems (13, 16).

During labour and delivery, few women were asked for permission before vaginal examinations. Similar trends in limited consent have been observed in other African studies, where autonomy during childbirth is often undermined (28). Privacy during vaginal examinations was similarly low. Moreover, there were disparities in access to pain relief during labour, with adolescents less likely to receive pain management in Ethiopia and Kenya compared to adults. This lower provision of pain relief for adolescents may reflect biases or insufficient provider training to address the specific needs of younger mothers (28–30).

Immediate postnatal care (PNC) before discharge was nearly optimal (80%) across all women and in Kenya and South Africa. In contrast, the rates were notably low in Ethiopia, particularly among adolescents, where only one-third received postnatal check-ups compared to over half of adult women. Moreover, adolescents received less postnatal counselling, particularly on topics like infant feeding, umbilical cord care, thermal care for newborns and maternal danger signs. PNC is critical for identifying and managing postpartum complications, particularly in the first 24 hours (31). Studies have shown that adolescents are less likely to seek PNC due to a lack of awareness, inadequate healthcare support, and cultural norms (16). This calls for interventions to increase the awareness of PNC and utilisation among adolescents.

One in five women reported experiencing mistreatment during childbirth, with higher rates among adolescents than adult women. These findings echo previous research highlighting the vulnerability of adolescents to mistreatment during childbirth due to age-related stigma and power imbalances in patient-provider interactions (28, 29, 32, 33). Privacy during delivery was reported at lower rates in Ethiopia, whereas in South Africa, adults were more likely to experience privacy than adolescents. The findings suggest systemic gaps in providing respectful maternity care to adolescents, which can deter them from seeking future care and exacerbate poor health outcomes (28, 29, 32). Despite these disparities, user satisfaction ratings for ANC and delivery care were comparable across age groups, with around 50% reporting positive experiences. Interestingly, in Ethiopia, adolescents were more likely than adults to recommend their childbirth provider, which may reflect lower expectations among younger women.

Obstetric complications, including prolonged labour and the need for blood transfusions, were significantly more common among adolescents, particularly in South Africa. Prolonged labour, possibly due to immature pelvic development and reduced uterine efficiency, increases risks of maternal exhaustion, obstetric fistula, and neonatal hypoxia (34, 35). High rates of blood transfusion suggest higher incidences of postpartum haemorrhage, often exacerbated by prolonged labour. These findings align with evidence linking adolescent pregnancies to higher risks of obstetric complications in resource-limited settings (2). These variations underscore the importance of strengthening emergency obstetric care and ensuring equitable access to skilled births by adolescents.

The findings revealed significant factors associated with both obstetric complications and mistreatment during delivery among adolescents. Adolescents from rural areas had higher odds of experiencing obstetric complications and mistreatment compared to their urban counterparts. This finding underscores the disparity in access to quality maternal healthcare services between rural and urban settings. Limited infrastructure, fewer skilled healthcare providers, and longer travel times to healthcare facilities in rural areas contribute to poorer outcomes and mistreatment (28, 30). This highlights the need for targeted interventions to ensure quality and respectful maternity care in rural settings

Antenatal depression emerged as a strong predictor of adverse obstetric outcomes and mistreatment. This highlights the critical role of mental health during pregnancy, as antenatal depression has been linked to poor maternal health behaviours, inadequate antenatal care utilisation, and biological stress responses affecting pregnancy outcomes (36, 37). Moreover, these factors illustrate the interplay between individual vulnerabilities and systemic failures. These findings call for integrating mental health screening and support into routine ANC programs and providing accessible psychosocial support services for pregnant adolescents.

Intimate partner violence (IPV) also emerged as a key risk factor for mistreatment during childbirth. IPV-exposed adolescents often experience increased vulnerability due to social stigma and inadequate support systems within maternal healthcare settings (38, 39). Addressing IPV within maternal healthcare requires a multisectoral approach, including training healthcare providers to screen for IPV, offering confidential counselling, and strengthening referral pathways to social support services.

Delivering in primary healthcare facilities was associated with lower odds of complications, suggesting the effectiveness of localised, community-level maternal care. This aligns with evidence from Campbell et al. (40), which emphasised the value of primary healthcare models in improving maternal health outcomes, particularly in LMICs. Similarly, adolescents from Kenya had significantly lower odds of mistreatment compared to those in Ethiopia. This regional variation may reflect differences in healthcare system structures, cultural practices, and implementation of respectful maternity care policies.

Government-operated facilities had higher mistreatment rates than private or NGO-run facilities, likely due to resource constraints and provider burnout (29). Investing in provider training and enforcing patient-centred care policies could help improve adolescents’ childbirth experiences. Similarly, fragmented care was another significant predictor of obstetric complications. Adolescents who visited multiple health facilities during labour faced a higher risk of complications, highlighting the detrimental impact of discontinuity in care. This underscores the need for a well-coordinated maternal healthcare system that provides consistent, high-quality care throughout pregnancy and childbirth. Strengthening referral systems and implementing integrated care models can help ensure seamless transitions between facilities, improving outcomes for adolescent mothers.

Finally, the perceptions of high-quality delivery care and privacy during delivery were protective factors against mistreatment. Adolescents who received privacy during delivery experienced significantly lower odds of mistreatment, aligning with other studies which demonstrated the importance of dignity and privacy in maternal care (41). These findings emphasise the need for healthcare systems to prioritise respectful care and ensure supportive environments to improve clinical maternal outcomes and experiences (41).

Despite these significant findings, this study is subject to some limitations. The reliance on self-reported experiences introduces potential biases, such as recall bias, particularly in the context of phone surveys. Adolescents may struggle to recall specific details of their healthcare experiences accurately, and social desirability bias could further influence their responses. Additionally, the small sample sizes in certain subgroups may limit the generalizability of the findings, particularly when comparing outcomes across diverse geographical and cultural contexts. While the longitudinal design provides a robust framework for analysing trends, it does not fully mitigate these challenges. Lastly, the focus on three countries limits the broader applicability of the findings to other LMICs with different healthcare settings.

## CONCLUSION

In conclusion, this study reports significant disparities in maternal healthcare access and quality among adolescents in Ethiopia, Kenya, and South Africa. Adolescents were less likely to initiate antenatal care early, attend the recommended number of visits, and receive essential maternal health interventions. However, it remains unclear whether these differences in care quality are entirely driven by the poorer socioeconomic status of younger pregnant women or whether discrimination in the health system and poorer patient activation leads to poorer care for these women. Additionally, adolescents faced higher rates of mistreatment during childbirth and an increased risk of obstetric complications. Risk factors such as rural residence, antenatal depression, and IPV increased their risks, highlighting the intersection of social, geographic, and mental health challenges in adolescent maternal healthcare.

Addressing these disparities requires a multi-faceted approach that strengthens adolescent-specific maternal health services, ensures respectful maternity care, and enhances emergency obstetric care. Policy interventions should prioritize integrating adolescent-friendly services into primary healthcare facilities alongside provider training in providing non-discriminatory care. Strengthening rural health infrastructure, incorporating mental health support into ANC, and implementing targeted interventions to address IPV are also essential. Future research should further explore the lived experiences of adolescent mothers to develop tailored, context-specific strategies that improve maternal health outcomes and promote equitable access to quality care.

## Supporting information

**S1 Table. Survey questions to assess mistreatment during facility-based childbirth in MNH Cohort Study (PDF)**

## Data Availability

All data used in this study data can be made available through a formal process of request to the KEMRI Institutional Data Access/Ethics Committee. The details of the guidelines can be found on the KEMRI Wellcome website (https://dataverse.harvard.edu/dataverse/kwtrp). Access to data can be provided via the KEMRI Wellcome Data Governance Committee.

## Acknowledgments

We would like to thank all participants for their time, effort, and contributions to the study.

## REFERENCES

1. Eyeberu A, Getachew T, Sertsu A, Sisay M, Baye Y, Debella A, et al. Teenage pregnancy and its predictors in Africa: A systematic review and meta-analysis. International Journal of Health Sciences. 2022;16(6):47.

2. World Health Organization. Adolescent pregnancy 2024 [Available from: https://www.who.int/news-room/fact-sheets/detail/adolescent-pregnancy.

3. World Health Organization. Adolescent health 2025 [Available from: https://www.who.int/health-topics/adolescent-health#tab=tab_1.

4. Akter M. Physical and psychological vulnerability of adolescents during pregnancy period as well as post traumatic stress and depression after child birth. Open Journal of Social Sciences. 2019;7(01):170.

5. Sully EA, Biddlecom A, Darroch JE, Riley T, Ashford LS, Lince-Deroche N, et al. Adding it up: investing in sexual and reproductive health 2019. 2020.

6. Kassa GM, Arowojolu A, Odukogbe A, Yalew AW. Prevalence and determinants of adolescent pregnancy in Africa: a systematic review and meta-analysis. Reproductive health. 2018;15:1–17.

7. Wado YD, Sully EA, Mumah JN. Pregnancy and early motherhood among adolescents in five East African countries: a multi-level analysis of risk and protective factors. BMC pregnancy and childbirth. 2019;19:1–11.

8. Carvajal L, Wilson E, Requejo JH, Newby H, de Carvalho Eriksson C, Liang M, et al. Basic maternal health care coverage among adolescents in 22 sub-Saharan African countries with high adolescent birth rate. Journal of global health. 2020;10(2).

9. Sabet F, Prost A, Rahmanian S, Al Qudah H, Cardoso MN, Carlin JB, et al. The forgotten girls: the state of evidence for health interventions for pregnant adolescents and their newborns in low-income and middle-income countries. The Lancet. 2023;402(10412):1580–96.

10. Chen X-K, Wen SW, Fleming N, Demissie K, Rhoads GG, Walker M. Teenage pregnancy and adverse birth outcomes: a large population based retrospective cohort study. International journal of epidemiology. 2007;36(2):368–73.

11. Honorato DJP, Fulone I, Silva MT, Lopes LC. Risks of adverse neonatal outcomes in early adolescent pregnancy using group prenatal care as a strategy for public health policies: a retrospective cohort study in Brazil. Frontiers in public health. 2021;9:536342.

12. Kumar M, Huang KY, Othieno C, Wamalwa D, Madeghe B, Osok J, et al. Adolescent Pregnancy and Challenges in Kenyan Context: Perspectives from Multiple Community Stakeholders. Glob Soc Welf. 2018;5(1):11–27.

13. Sidamo NB, Kerbo AA, Gidebo KD, Wado YD. Socio-ecological analysis of barriers to access and utilization of adolescent sexual and reproductive health services in sub-saharan africa: a qualitative systematic review. Open Access Journal of Contraception. 2023:103–18.

14. Maheshwari MV, Khalid N, Patel PD, Alghareeb R, Hussain A. Maternal and neonatal outcomes of adolescent pregnancy: a narrative review. Cureus. 2022;14(6).

15. Thirukumar M, Thadchanamoorthy V, Dayasiri K. Adolescent Pregnancy and Outcomes: A Hospital-Based Comparative Study at a Tertiary Care Unit in Eastern Province, Sri Lanka. Cureus. 2020;12(12):e12081.

16. Tolossa T, Gold L, Dheresa M, Turi E, Yeshitila Y, Abimanyi-Ochom J. Adolescent maternal health services utilization and associated barriers in Sub-Saharan Africa: A comprehensive systematic review and meta-analysis before and during the sustainable development goals. Heliyon. 2024.

17. Mekonnen T, Dune T, Perz J. Maternal health service utilisation of adolescent women in sub-Saharan Africa: a systematic scoping review. BMC pregnancy and childbirth. 2019;19:1–16.

18. Kayemba V, Kabagenyi A, Ndugga P, Wasswa R, Waiswa P. Timing and quality of antenatal care among adolescent mothers in a rural community, Uganda. Adolescent health, medicine and therapeutics. 2023:45–61.

19. Arsenault C, Mfeka-Nkabinde NG, Chaudhry M, Jarhyan P, Taddele T, Mugenya I, et al. Antenatal care quality and detection of risk among pregnant women: An observational study in Ethiopia, India, Kenya, and South Africa. PLoS Med. 2024;21(8):e1004446.

20. Wright K, Mugenya I, Clarke-Deelder E, Baensch L, Taddele T, Mebratie AD, et al. Implementation of Maternal and Newborn Health Mobile Phone E-Cohorts to Track Longitudinal Care Quality in Low-and Middle-Income Countries. Global Health: Science and Practice. 2024;12(4).

21. Leslie HH, Sharma J, Mehrtash H, Berger BO, Irinyenikan TA, Balde MD, et al. Women’s report of mistreatment during facility-based childbirth: validity and reliability of community survey measures. BMJ Global Health. 2022;5(Suppl 2):e004822.

22. Kapula N, Sacks E, Wang DT, Odiase O, Requejo J, Afulani PA. Associations between self-reported obstetric complications and experience of care: a secondary analysis of survey data from Ghana, Kenya, and India. Reproductive health. 2023;20(1):7.

23. Yaya S, Bishwajit G, Ekholuenetale M, Shah V, Kadio B, Udenigwe O. Timing and adequate attendance of antenatal care visits among women in Ethiopia. PloS one. 2017;12(9):e0184934.

24. Pell C, Menaca A, Were F, Afrah NA, Chatio S, Manda-Taylor L, et al. Factors affecting antenatal care attendance: results from qualitative studies in Ghana, Kenya and Malawi. PloS one. 2013;8(1):e53747.

25. Adde KS, Dickson KS, Amu H. Prevalence and determinants of the place of delivery among reproductive age women in sub–Saharan Africa. Plos one. 2020;15(12):e0244875.

26. Shiferaw S, Spigt M, Godefrooij M, Melkamu Y, Tekie M. Why do women prefer home births in Ethiopia? BMC pregnancy and childbirth. 2013;13:1–10.

27. Tamiso A, Jisso M, Abera N, Alemayehu A, Gadisa A, Umer A, et al. Barriers Towards Obstetric Care Service Utilization in Ethiopia: An Explorative Qualitative Study. Ethiopian Journal of Health Sciences. 2023;33(2).

28. Bohren MA, Vogel JP, Hunter EC, Lutsiv O, Makh SK, Souza JP, et al. The mistreatment of women during childbirth in health facilities globally: a mixed-methods systematic review. PLoS medicine. 2015;12(6):e1001847.

29. Freedman LP, Ramsey K, Abuya T, Bellows B, Ndwiga C, Warren CE, et al. Defining disrespect and abuse of women in childbirth: a research, policy and rights agenda. Bull World Health Organ. 2014;92(12):915–7.

30. Kruk ME, Kujawski S, Moyer CA, Adanu RM, Afsana K, Cohen J, et al. Next generation maternal health: external shocks and health-system innovations. The Lancet. 2016;388(10057):2296–306.

31. Organization WH. WHO recommendations on postnatal care of the mother and newborn: World Health Organization; 2014.

32. Ajayi AI, Gebrekristos LT, Otukpa E, Kabiru CW. Adolescents’ experience of mistreatment and abuse during childbirth: a cross-sectional community survey in a low-income informal settlement in Nairobi, Kenya. BMJ Global Health. 2023;8(11):e013268.

33. Irinyenikan TA, Aderoba AK, Fawole O, Adeyanju O, Mehrtash H, Adu-Bonsaffoh K, et al. Adolescent experiences of mistreatment during childbirth in health facilities: secondary analysis of a community-based survey in four countries. BMJ global health. 2022;5(Suppl 2):e007954.

34. Ganchimeg T, Ota E, Morisaki N, Laopaiboon M, Lumbiganon P, Zhang J, et al. Pregnancy and childbirth outcomes among adolescent mothers: a World Health Organization multicountry study. Bjog. 2014;121 Suppl 1:40–8.

35. Neal S, Channon AA, Chandra-Mouli V, Madise N. Trends in adolescent first births in sub-Saharan Africa: a tale of increasing inequity? International journal for equity in health. 2020;19:1–11.

36. Wainaina CW, Sidze EM, Maina BW, Badillo-Amberg I, Anyango HO, Kathoka F, et al. Psychosocial challenges and individual strategies for coping with mental stress among pregnant and postpartum adolescents in Nairobi informal settlements: a qualitative investigation. BMC pregnancy and childbirth. 2021;21:1–11.

37. Grote NK, Bridge JA, Gavin AR, Melville JL, Iyengar S, Katon WJ. A meta-analysis of depression during pregnancy and the risk of preterm birth, low birth weight, and intrauterine growth restriction. Archives of general psychiatry. 2010;67(10):1012–24.

38. White SJ, Sin J, Sweeney A, Salisbury T, Wahlich C, Montesinos Guevara CM, et al. Global prevalence and mental health outcomes of intimate partner violence among women: A systematic review and meta-analysis. Trauma, Violence, & Abuse. 2024;25(1):494–511.

39. Chambliss LR. Intimate partner violence and its implication for pregnancy. Clin Obstet Gynecol. 2008;51(2):385–97.

40. Campbell OM, Graham WJ. Strategies for reducing maternal mortality: getting on with what works. The lancet. 2006;368(9543):1284–99.

41. Sacks E, Kinney MV. Respectful maternal and newborn care: building a common agenda. Reproductive health. 2015;12(1):1–4.

